# OpenSAFELY NHS Service Restoration Observatory 2: changes in primary care activity across six clinical areas during the COVID-19 pandemic

**DOI:** 10.1101/2022.06.01.22275674

**Authors:** Helen J Curtis, Brian MacKenna, Milan Wiedemann, Louis Fisher, Richard Croker, Caroline E Morton, Peter Inglesby, Alex J Walker, Jessica Morley, Amir Mehrkar, Sebastian CJ Bacon, George Hickman, David Evans, Tom Ward, Simon Davy, William J Hulme, Orla Macdonald, Robin Conibere, Tom Lewis, Martin Myers, Shamila Wanninayake, Kiren Collison, Charles Drury, Miriam Samuel, Harpreet Sood, Andrea Cipriani, Seena Fazel, Manuj Sharma, Wasim Baqir, Chris Bates, John Parry, Ben Goldacre

## Abstract

**Background:** The COVID-19 pandemic has disrupted healthcare activity across a broad range of clinical services. The NHS stopped non-urgent work in March 2020, later recommending services be restored to near-normal levels before winter where possible.

**Aims:** Using routinely collected data, our aim was to describe changes in the volume and variation of coded clinical activity in general practice in: (i) cardiovascular disease, (ii) diabetes, (iii) mental health, (iv) female and reproductive health, (v) screening, and (vi) processes related to medication.

**Design and setting:** With the approval of NHS England, we conducted a cohort study of 23.8 million patient records in general practice, in-situ using OpenSAFELY.

**Methods:** We selected common primary care activity using CTV3 codes and keyword searches from January 2019 - December 2020, presenting median and deciles of code usage across practices per month.

**Results:** We identified substantial and widespread changes in clinical activity in primary care since the onset of the COVID-19 pandemic, with generally good recovery by December 2020. A few exceptions showed poor recovery and warrant further investigation, such as mental health, e.g. “Depression interim review” (median across practices in December 2020 -41.6% compared to December 2019).

**Conclusions:** Granular NHS GP data at population-scale can be used to monitor disruptions to healthcare services and guide the development of mitigation strategies. The authors are now developing real-time monitoring dashboards for key measures identified here as well as further studies, using primary care data to monitor and mitigate the indirect health impacts of Covid-19 on the NHS.

**How this fits in:** During the COVID-19 pandemic, routine healthcare services in England faced significant disruption, and NHS England recommended restoring NHS services to near-normal levels before winter 2020. Our previous report covered the disruption and recovery in pathology tests and respiratory activity: here we describe an additional six areas of common primary care activity. We found most activities exhibited significant reductions during pandemic wave 1 (with most recovering to near-normal levels by December); however many important aspects of care - especially those of a more time-critical nature - were maintained throughout the pandemic. We recommend key measures for ongoing monitoring and further investigation of the impacts on health inequalities, to help measure and mitigate the ongoing indirect health impacts of COVID-19 on the NHS.

## Background

The COVID-19 pandemic disrupted healthcare services globally.^1^ In March 2020, NHS England initially promoted measures to reduce viral transmission and provide only essential health services.^2,3^ The World Health Organisation (WHO) recommended rapid assessments of healthcare capacity and the development of key performance indicators.^4,5^ From August 2020, NHS England aimed to restore primary care and other services back to normal activity where clinically appropriate.^6^

Various studies have assessed the impact of the pandemic on non-COVID health services, for example mental health, female and reproductive health, health screenings, and prescribing.^7–10^ Overall, findings suggest a substantial decrease from the first official lockdown in the UK (March 2020), with recovery to near-normal activity in most clinical areas from around July 2020. We previously reported a data-driven approach for monitoring healthcare disruptions and recovery using the OpenSAFELY platform (https://www.opensafely.org/) combined with input from a clinical advisory group, intended to explore changes in high volume areas, including those that might otherwise be missed.^11^ Selecting pathology tests and respiratory conditions as key examples, we showed that activity largely decreased substantially and subsequently recovered. However, we also found that some activities such as blood coagulation tests were well-maintained, suggesting that important clinical care was effectively prioritised.^11^ Certain clinical conditions like cardiovascular disease (CVD) and diabetes are associated with higher risk of morbidity and mortality from COVID-19, emphasising the importance of maintaining good routine care.^12–15^

We therefore set out to generate an overall picture of changes in clinical activity in primary care across key areas of medicine; and to identify key measures to continuously monitor the impacts of COVID-19 on the NHS and inform further studies. Specifically we extended our earlier study^11^ to December 2020 and expanded our work to cover six further clinical areas: cardiovascular disease, diabetes, mental health, female and reproductive health, screening procedures, and processes related to medication.

## Methods

### Study design and data source

Following the methodology previously described^11^, we conducted a cohort study using routinely collected pseudonymised primary care electronic health records (EHR) in the OpenSAFELY-TPP platform (https://opensafely.org). This included 40% of England’s general practices (2,500, those using TPP SystmOne software), covering 24m patients. In brief, this structured GP data includes one record for every diagnostic code, prescription, blood test, investigation, or similar in primary care.

### Study population and data processing

The study population and data processing were as in our previous study^11^, except for a later study end date (31st December 2020), and included all patients registered with a general practice on 31st December 2020. Briefly, we counted occurrences of all coded clinical activities per month throughout 2019-2020, by Clinical Terms Version 3 (CTV3) code (including diagnoses, investigations and other clinical/administrative processes, but excluding medications/vaccinations), grouped by general practice. We excluded any codes with less than 1000 total occurrences in 2020 to identify the most frequent clinical activities. We used the CTV3 parent-child hierarchy (e.g. 24: *‘Examination of cardiovascular system’*; 242: ‘*O/E - pulse rate’*) to group similar activities, and mapped each code to high level CTV3 concepts to assist with categorisation into broad topics (e.g. “cardiovascular”). For each code/group we calculated the monthly rate of code usage per 1000 registered patients and the median and deciles across practices.

### Study measures

We pragmatically grouped activity and selected clinical codes relevant to each of the following topics: Cardiovascular disease, Diabetes, Mental health, Female and reproductive health, Screening and related procedures, and Processes related to medication. We did not include prescribing, which is coded in GP systems using the NHS dictionary of medicines and devices, as high quality routinely updated analysis of primary care dispensing data is already openly available on our partner service OpenPrescribing (https://openprescribing.net/). The selection of clinical codes was largely based on existing CTV3 concepts and keyword searching. A detailed description of our methodology is available in Supplementary Information and Table S1.

### Clinical advisory group

We established a clinical advisory group to review our findings, consisting of general practitioners, pharmacists, pathologists, other relevant specialists, and national clinical advisors, formed by invitation of clinicians known through existing professional relationships. The raw results of our data driven approach on each topic were discussed with the advisory group during a series of online meetings to prioritise clinical topics and inform interpretation. The group also had the opportunity to comment on these documents outside of the meetings, and if needed arrange further meetings with the research team. Additionally the group was asked to select “key measures” of activity from each clinical area to direct targeted work on health inequalities.

### Software and reproducibility

Data management and analysis were performed using Python 3.8. Code for data management and analysis are available online (https://github.com/opensafely/restoration-observatory-data-driven).

### Patient and public involvement

We have developed a publicly available website https://opensafely.org/ through which we invite any patient or member of the public to contact us regarding this study or the broader OpenSAFELY project.

## Results

All included clinical codes/groups for each topic along with their total 2020 usage are shown in Tables S2-S7. We summarise results by topic, highlighting selected code usage at key points in the pandemic in England (February, April and December 2020; Table 1).

**Table 1.**
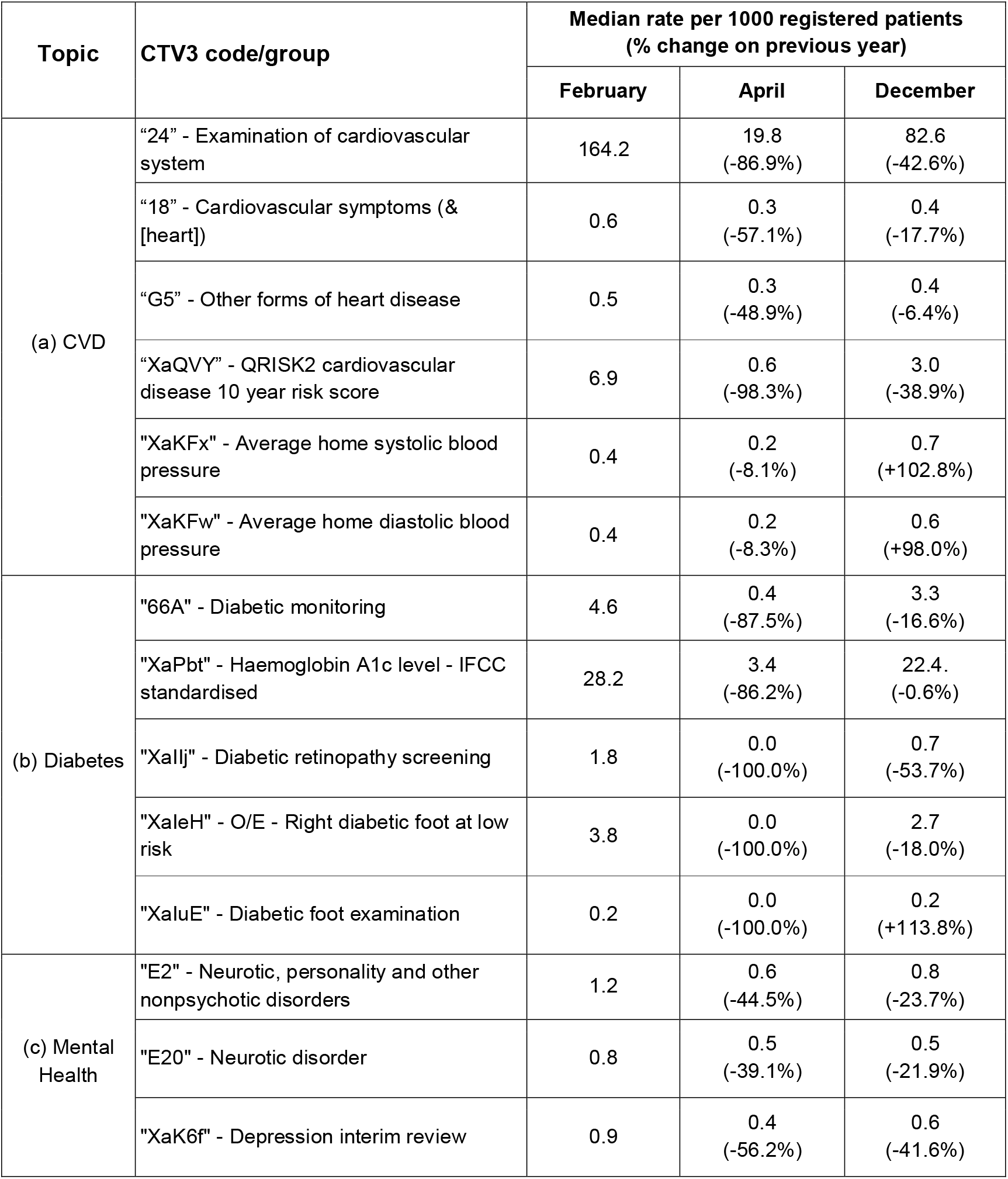
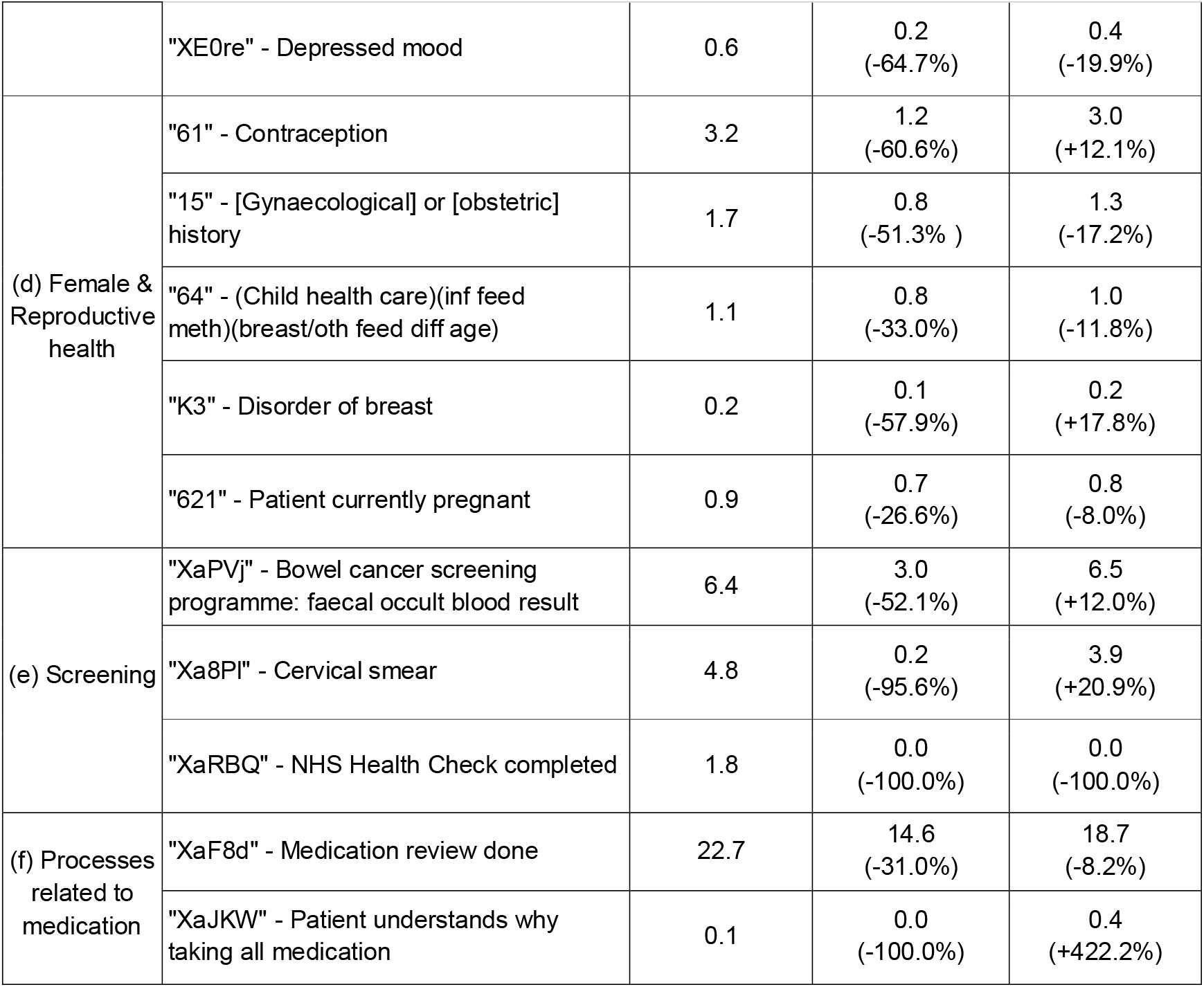
Rate of recording of selected CTV3 codes and groups of codes for each topic in February, April and December 2020. (a) cardiovascular disease (CVD), (b) diabetes, (c) mental health, (d) female & reproductive health, (e) screening procedures, and (f) processes related to medication. Showing median rate across English practices per 1000 registered patients and % change on same month in previous year. Further detailed breakdowns of code recording are shown in Table S2.

### Cardiovascular disease (CVD)

The majority of cardiovascular disease coded activity experienced a substantial decline during the initial stages of the pandemic, with limited recovery by September 2020 that levelled off through to December 2020, e.g. blood pressure recording and electrocardiography (Figure 1a-b).

**Figure 1.**
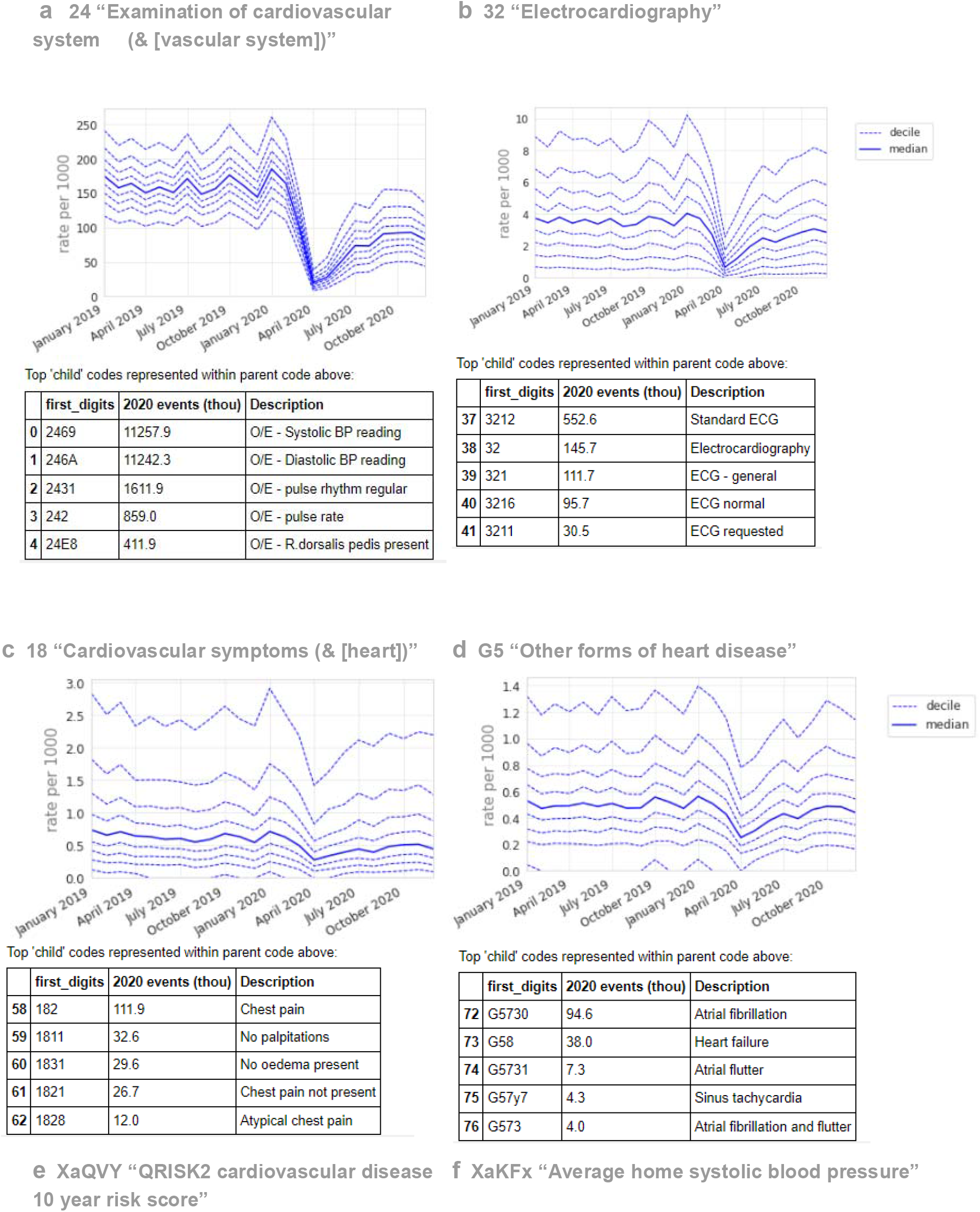
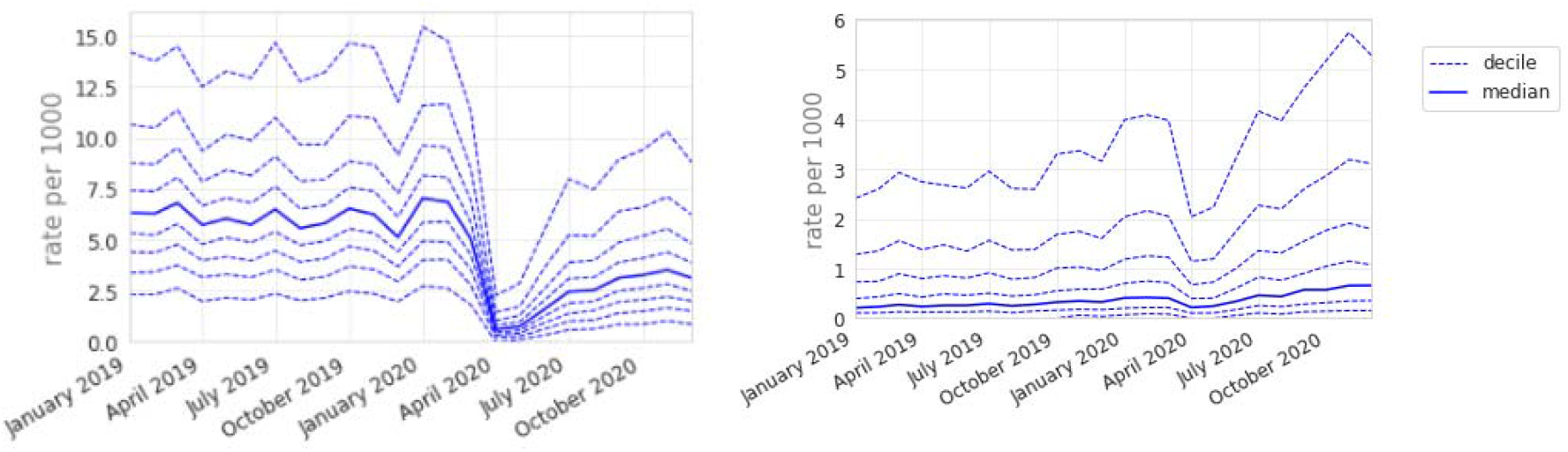
Recording of grouped subsets of cardiovascular CTV3 codes across TPP practices in England (January 2019 - December 2020). (a) “Examination of cardiovascular system (& [vascular system])”. (b) “Electrocardiography”, (c) “Cardiovascular symptoms (& [heart])”, (d) “Other forms of heart disease”, (e) “QRISK2 cardiovascular disease 10 year risk score”, (f) “Average home systolic blood pressure”. Each group includes CTV3 codes that begin with the digits shown and is not necessarily an exhaustive collection of every activity related to the description. For grouped codes (a-d), the top 5 codes represented within each group are listed in tables below each graph.

Exceptions included symptoms related to cardiovascular system (Figure 1c), which experienced a sustained drop (April -57.1%, December -17.7%); “other forms of heart disease” (Figure 1d), including atrial fibrillation and heart failure, experienced a small drop but largely recovered (April -48.9%, December -6.4%); QRISK2 (XaQVY), a CVD risk tool, dropped dramatically (−98.3%) with limited recovery (December -38.9%). Blood pressure at home codes increased overall (+100% in December, Figure 1f), but were infrequently used (410k, 400k total events, Table S2) compared to blood pressure codes recorded with no setting (11 million, Figure 1a).

### Diabetes

Diabetic monitoring and HbA1c testing both experienced a large drop with good recovery (April -87.5%, December -16.6%; April -86.2%, December -0.6%; Table 1b, Figure 2a-b). For Diabetic Retinopathy Screening, the median dropped to zero in April, with limited recovery by December (−53.7%; Figure 2c). For “right diabetic foot at low risk”, the median dropped to zero in April, with good recovery by December (−18.0%; Figure 2d). There was substantial variation in the rate of diabetes monitoring and retinopathy screening at baseline, indicating some incompleteness.

**Figure 2.**
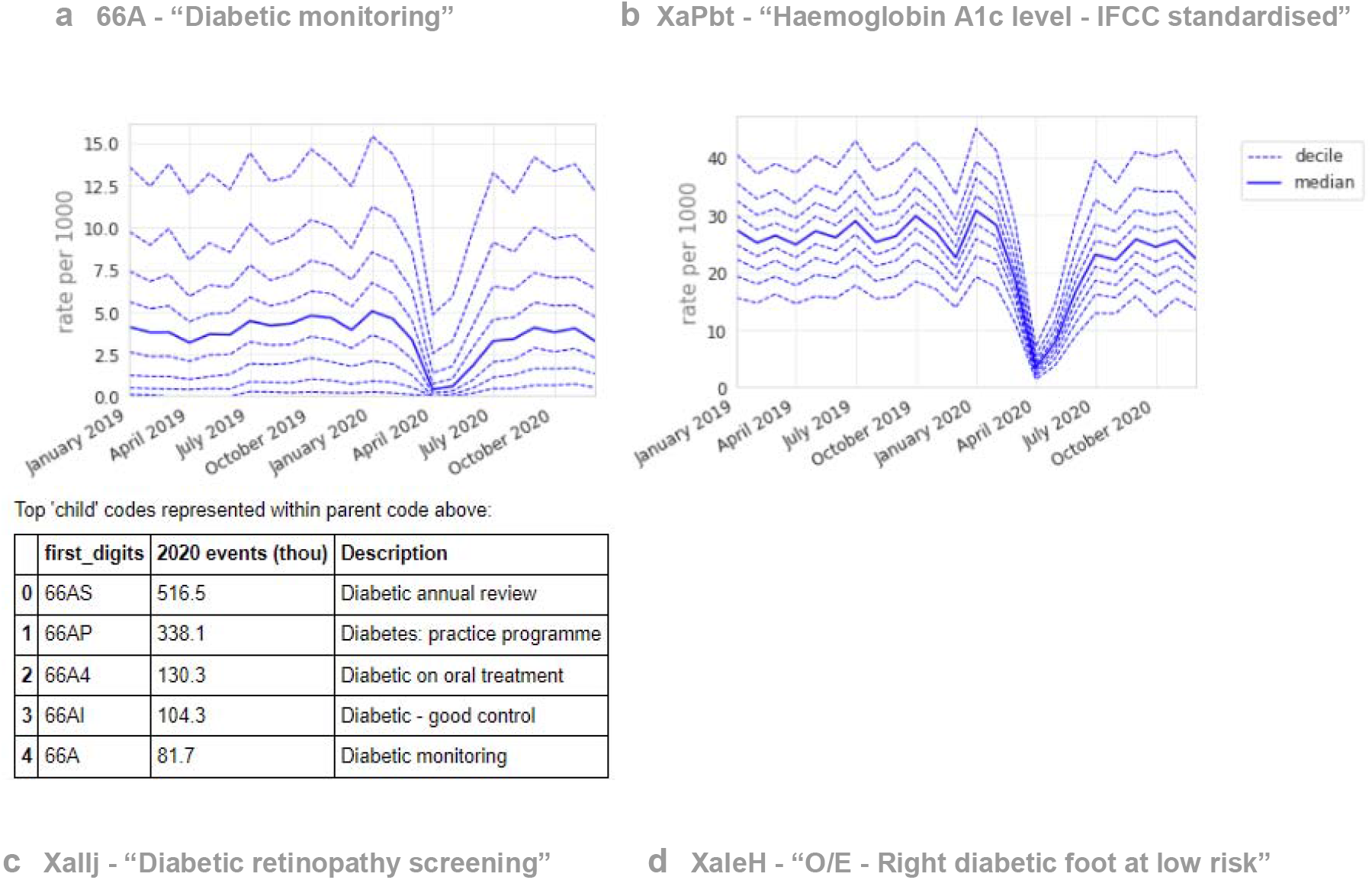
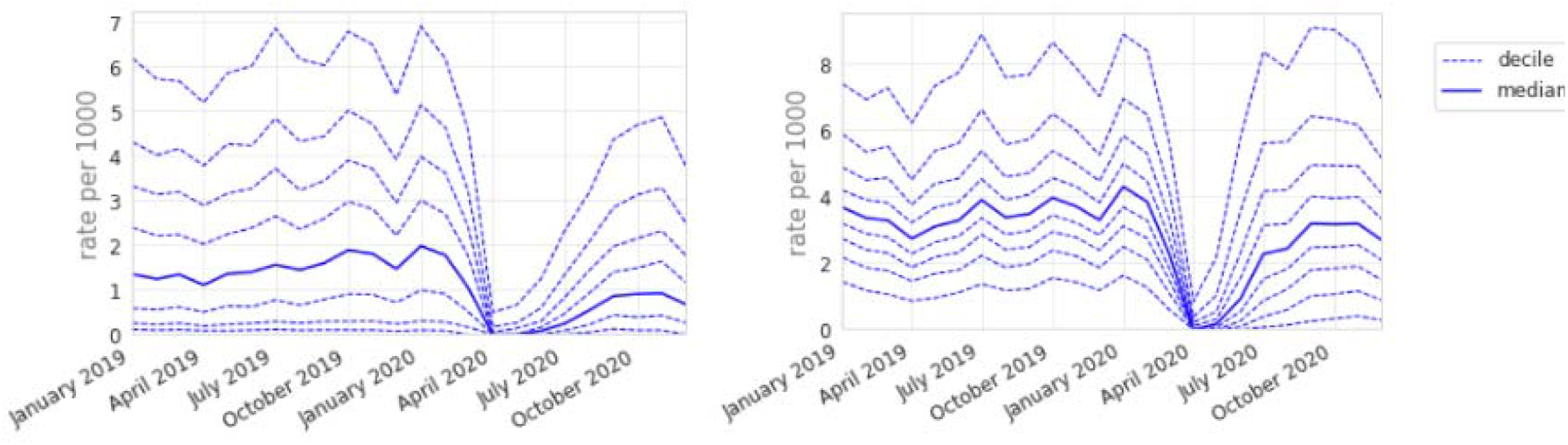
Recording of CTV3 codes across TPP practices in England (January 2019 - December 2020). (a) “Diabetic monitoring”, (b) “Haemoglobin A1c level - IFCC standardised”, (c) “Diabetic retinopathy screening”, (d) “O/E - Right diabetic foot at low risk”. Each code is not necessarily an exhaustive collection of every activity related to the description. O/E = “on examination”. “O/E - Left diabetic foot at low risk” is not shown but has an almost identical pattern to the right foot equivalent. Only ‘at low risk’ codes were included as codes related to moderate or increased risk did not meet the frequency threshold for the data driven analysis. For grouped codes (a), the top 5 codes represented within the group are listed under the graph.

### Mental health

The majority of mental health coded activity experienced a moderate decline during the initial stages of the pandemic, with incomplete recovery by December 2020 (Table 1c), e.g. “Neurotic, personality and other nonpsychotic disorders”: April -44.5%, December -23.7% (Figure 3a); “Depressed mood”: April -64.7%, December -19.9% (Figure 3b). “Depression interim review” activity showed particularly poor recovery (April -56.2%, December -41.6%, Figure 3c).

**Figure 3.**
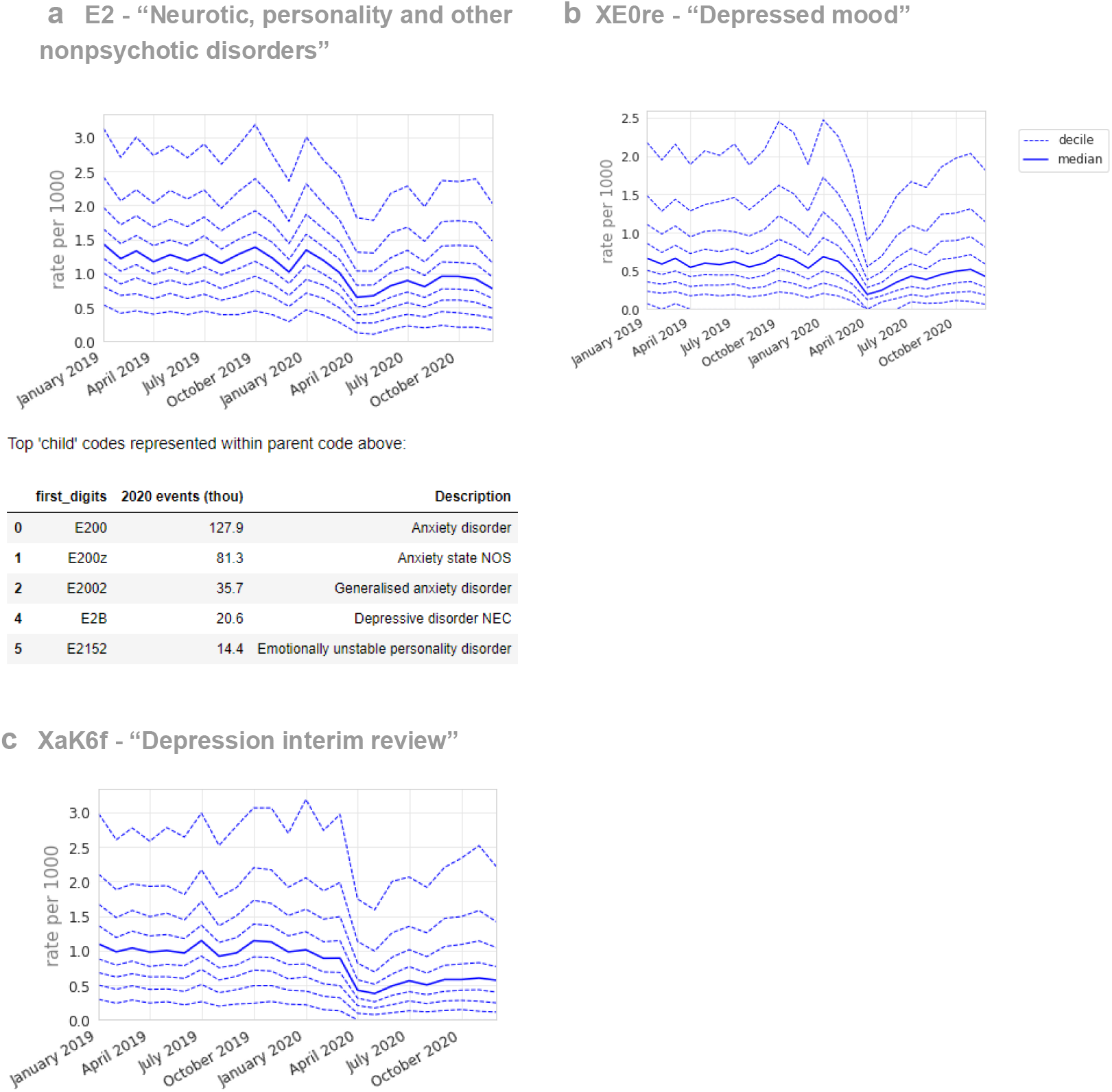
Recording of CTV3 codes across TPP practices in England (January 2019 - December 2020). (a) “Neurotic, personality and other nonpsychotic disorders”, (b) “Depression interim review”, (c) “Depressed mood”. Each code is not necessarily an exhaustive collection of every activity related to the description. For grouped codes (a), the top 5 codes represented within the group are listed under the graph.

### Female and reproductive health

The majority of female and reproductive-related activity experienced a moderate decline during the initial stages of the pandemic, with either recovery to near-normal levels by December 2020 or a return to an existing increasing or decreasing trend; for example “Contraception” (codes beginning “61”): April -60.6%, December +12.1% (Figure 4a).

**Figure 4.**
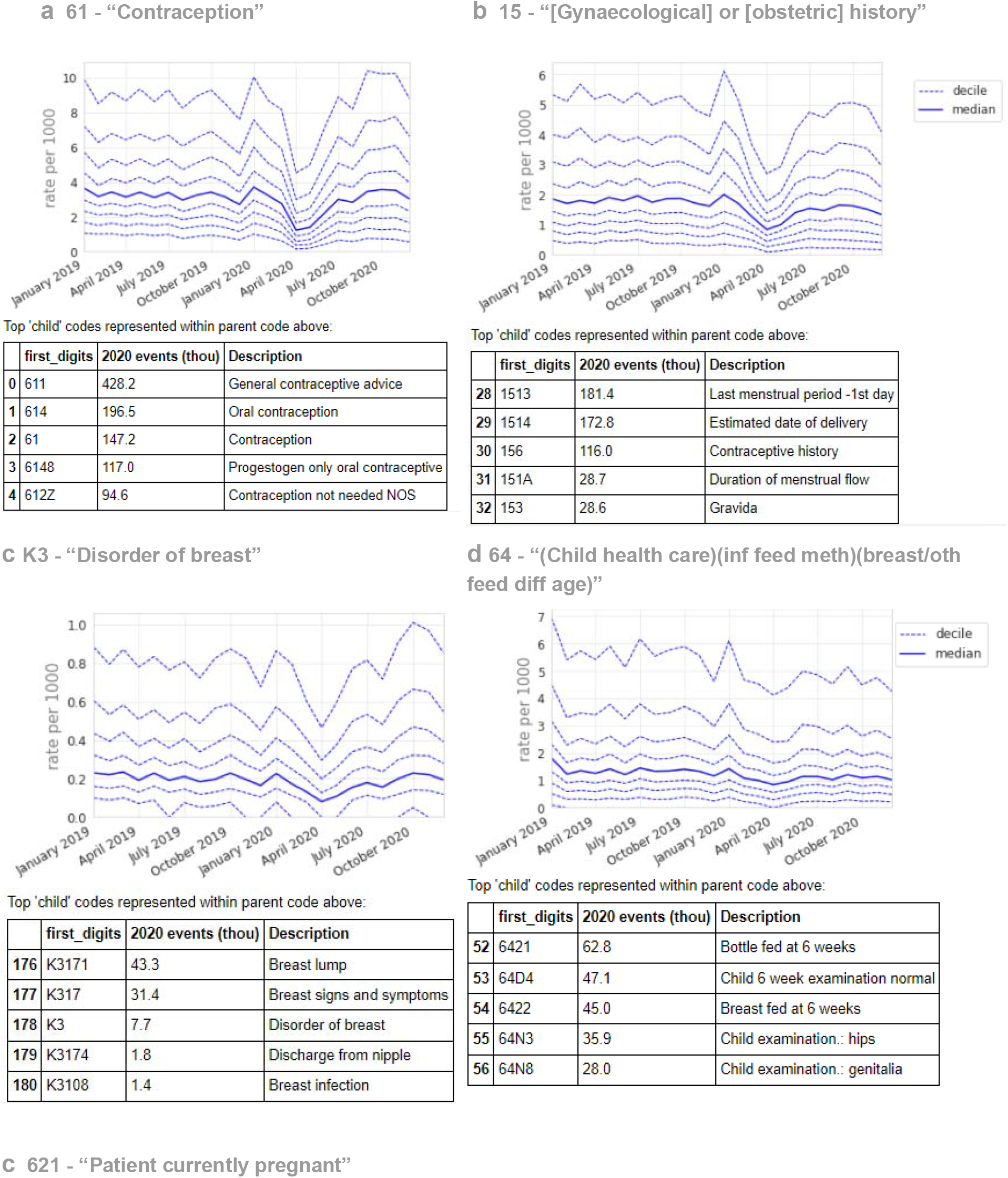
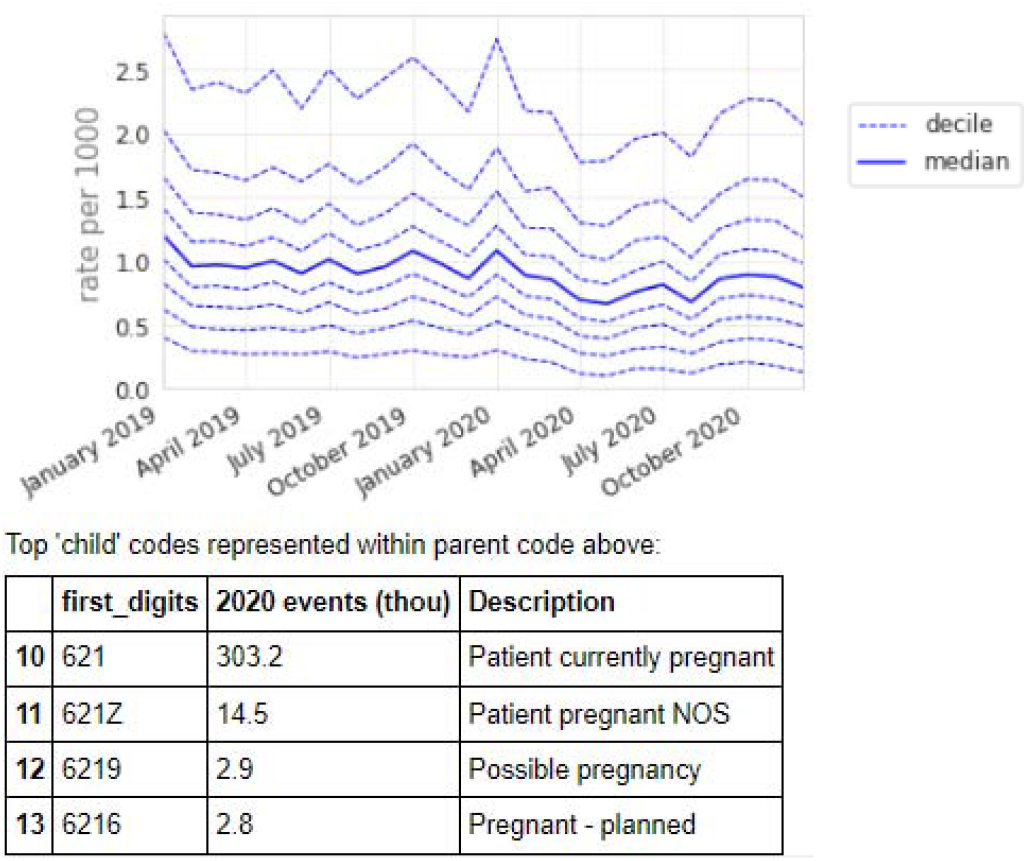
Recording of grouped subsets of female and reproductive health-related CTV3 codes across TPP practices in England (January 2019 - December 2020). (a) “Contraception”. (b) “[Gynaecological] or [obstetric] history”, (c) “Patient currently pregnant”. Each CTV3 code does not necessarily represent all activity related to the description. The top 5 codes represented within each group are listed under the graph.

“[Gynaecological] or [obstetric] history” showed a broadly similar pattern but with a slight sustained reduction (December -17.2%, Figure 4b). Various codes encompassed gynaecological and obstetric procedures and symptoms, which generally recovered to pre-pandemic levels, although many had a median of zero throughout the period. One example was “Disorder of breast” (Figure 4c, the most common code within which was “Breast lump”), which had a small increase by December (April -57.9%; December +17.8%).

Codes related to infant health such as six-week checks only had a small drop and recovered to near-normal levels, perhaps following a slightly decreasing trend (Figure 4d).

Codes for “Patient currently pregnant” reduced slightly and a small reduction was maintained through to December (Figure 4e).

### Screening and related activity

The majority of screening and related activity experienced a substantial decline, with recovery to slightly above normal levels by December 2020 (Table 1e), e.g. “Bowel cancer screening programme: faecal occult blood result” (April median -52.1%, December +12.0%) and “Cervical screening” (April -95.6%, December +20.9%). However, the patterns of recovery differed, with bowel cancer screening only beginning to recover around September while cervical screening was near normal by July (Figure 5a-b).

**Figure 5.**
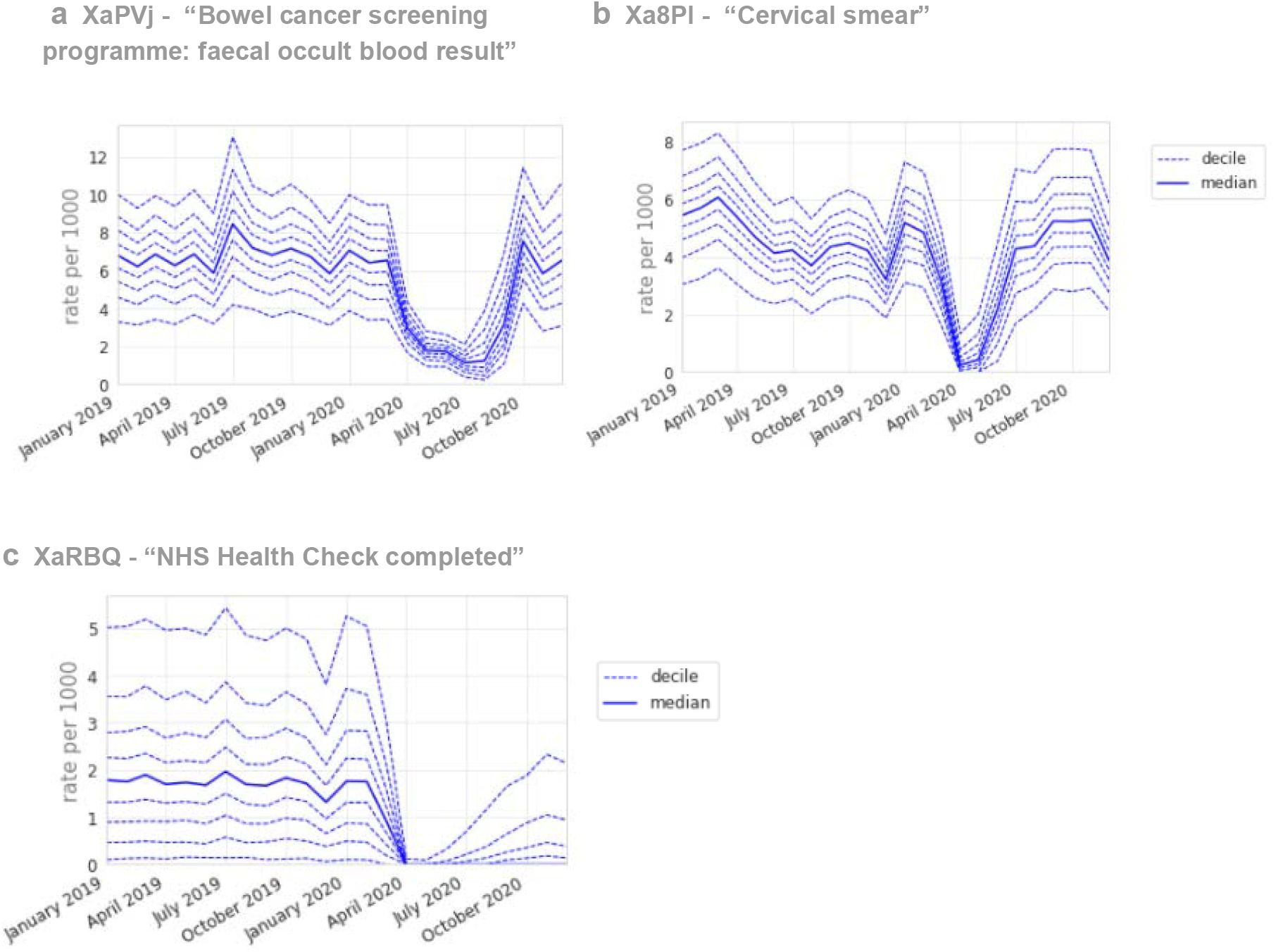
Recording of screening-related CTV3 codes across TPP practices in England (January 2019 - December 2020). (a) “Bowel cancer screening programme: faecal occult blood result”. (b) “Cervical Screening”, (c) “NHS Health Check completed”. Each CTV3 code does not necessarily represent all activity related to the description. Note, Diabetic retinopathy screening was also included with screening codes, but is discussed in the Diabetes section.

NHS health checks reduced from 1.8 per thousand in February to close to zero activity in April 2020 (median 0.0, with some recovery but median remaining zero by December 2020 (Figure 5c). Alcohol screening had a skewed distribution with a median of only 0.1 records per thousand in February 2020, reducing to zero in April and December 2020.

### Processes related to medication

Several codes explicitly mentioned “medication review”, most commonly XaF8d “Medication review done” (7.25m occurrences, Figure 6a), which experienced a small dip and gradual recovery (April median -31.0%, December -8.2%). Other medication review codes (e.g. review by pharmacist, for a specific disease, or indicating presence/absence of patient), were used less consistently between practices.

**Figure 6.**
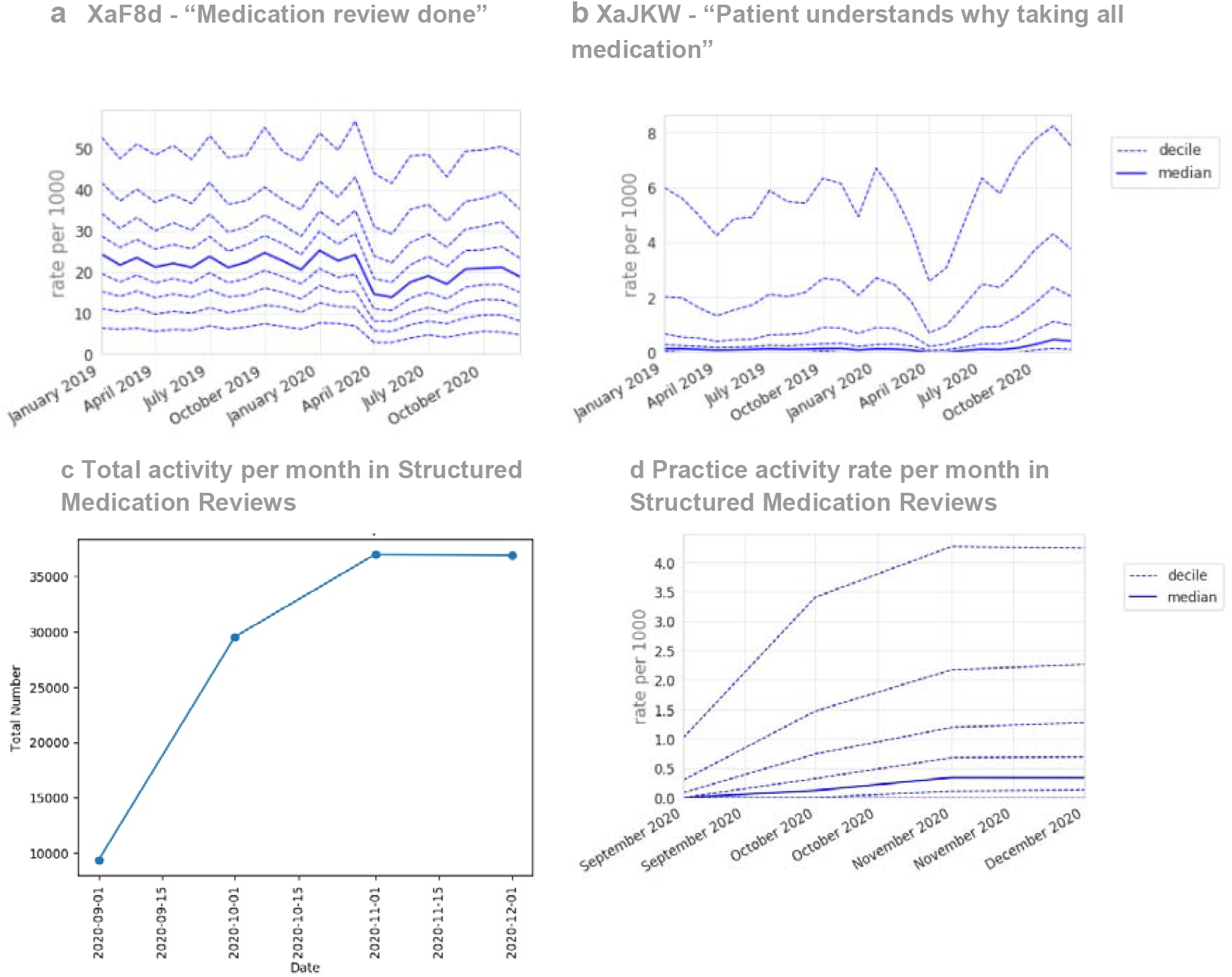
Recording of CTV3 codes across TPP practices in England. (a) “Medication review done” (January 2019 - December 2020) (b) “Patient understands why taking all medication”.(c) Total recording of Structured Medication Reviews across TPP practices in England throughout their period of use to date (September 2020 - December 2020), and (d) practice deciles showing the rate per 1000 registered patients for Structured Medication Reviews. Each code is not necessarily an exhaustive collection of every activity related to the description.

Several codes, generally uncommon but increasing in usage, were likely recorded as part of a medication review, such as “Patient understands why taking all medication” (Figure 6b; February median 0.1, April 0.0, December 0.4), and “Able to manage medication” (Xa2yC).

From September 2020 Structured medication reviews appeared (Figure 6c) and increased rapidly to around 350k records per month; however, some practices were recording them at much higher rates than others (Figure 6d).

## Discussion

### Summary

We identified widespread but also heterogeneous changes in clinical activity in primary care since the onset of the COVID-19 pandemic. There was generally good recovery by December 2020, with some exceptions: notably mental health, which showed minimal recovery. There was also variation from the median across practices, for both baseline and recovery.

### Strengths and weaknesses

The key strengths are the scale and completeness of the underlying raw EHR data, available close to real time, and we engaged with clinicians for added context. All processed data and analytical code is openly available in the Supplementary Materials or Github. We will publish our recommended key measures in a live updating report, and we encourage other groups to use OpenSafely for further exploration. Our data-driven approach is intended to generate an overall picture of primary care clinical activity, and explore high volume areas that might otherwise be missed, for example when not included in manually curated codelists.

Despite the strengths we recognise some limitations as previously discussed.^11^ Our data-driven approach and filtering processes may have omitted some relevant codes; codes do not necessarily indicate unique or new events, and may be affected by changes in coding behaviour. All coded activity for patients registered at the end of the study period were included, and all activity was included under their latest practice. Patients who died or deregistered from TPP practices during the study period were not included. Overall, activity counts were up to 6-8% lower than database totals in the earliest months of the study period.

### Interpretation and context for each clinical area

Given the diversity of clinical areas covered by this overarching analysis, the clinical advisory group evaluated and interpreted the variation for each clinical area separately.

#### CVD

Much coded activity related to monitoring, and remained around 40% reduced from pre-pandemic levels. This was not surprising due to changes in guidance and financial incentives.^16^ We warn that electrocardiogram data should be interpreted cautiously as this is often conducted outside primary care and not always systematically coded. The lack of recovery in QRISK2 scores may have public health significance, potentially causing later diagnosis of heart disease and poorer early management. Home blood pressure coding unsurprisingly increased, however home monitoring in general may not always be recorded completely or consistently in GP records. The consistent pattern of decrease in most cardiovascular related activity is in line with results from other studies in the UK.^8,17,18^ This is of particular interest because delays in the management of high blood pressure are associated with worse clinical outcomes.^19^ The clinical advisory group proposed “blood pressure monitoring” and “QRISK2 risk scores” (or any cardiovascular risk score codes including the newer QRISK3 codes) as key measures.

#### Diabetes

Routine diabetes care almost entirely stopped, but largely rapidly recovered, e.g. HbA1c testing was near normal by December 2020. Diabetes monitoring and foot checks remained slightly below normal. In some areas, concerning foot changes may be seen by specialist services, hence may sometimes appear reduced in primary care. Diabetic retinopathy screening recovered less well; however, specialist clinics conduct this service and send reports to primary care which are manually coded, therefore the sustained drop may indicate a coding change or other provider changes. Most diabetes care activity varied widely between practices, possibly due to differences in demographics and prevalence; coding (e.g. use of data entry templates in electronic health record systems); use of external providers; or quality of care. Our results add to the findings from earlier studies which reported a rapid decline in the rate of new diabetes mellitus (DM) diagnoses and HbA1c testing in April 2020,^8,18^ and are in line with data showing good but incomplete recovery in the following months.^20,21^ The clinical advisory group proposed HbA1c testing as a key sentinel measure.

#### Mental Health

Most mental health activity coded by GPs showed a sustained reduction. This was consistent across various markers of activity. This was surprising given the much-discussed impact of the pandemic on mental health^22,23^, but may be explained by patients either not seeking help or choosing other services/online resources. The latter are unlikely to explain all the reduction. For example, a recent study on mental health and telepsychiatry showed that the rapid shift to remote service delivery has not reached some groups of patients (in particular patients with dementia and mild cognitive impairment) who may require more tailored management^24^. Dementia was not widely represented in our results, perhaps being covered by a range of CTv3 codes; we will conduct further research on the impacts of COVID on dementia in primary care to capture this fully.

The reduction in “Depression interim review” may warrant further investigation, but could reflect a change in coding behaviour. However, the similar reduction in codes for depressed mood would argue against this as the sole explanation. Nationally, the prescribing of antidepressants in primary care was sustained, indicating that access to some treatment was maintained (Figure S1). Further analyses are planned before proposing any single measure for immediate ongoing monitoring, as mental health activity (especially for depression and other mood disorders) spans different services such as community mental health trusts^25^, which have limited coverage in OpenSAFELY.

Previous research similarly showed that primary care-recorded diagnosis of common mental health conditions, and associated prescribing, reduced significantly in early 2020 and did not recover to pre-pandemic levels by the end of 2020.^8,18,26^ One region found a reduction in self harm in primary care sustained through to May 2021.^27^ Other studies suggest that the impact on mental health may have been temporary, but following a generally worsening trend^28^ and the English health department have responded by developing a targeted action plan.^29^

#### Female and Reproductive Health

Female and reproductive health clinical activity generally declined modestly around April 2020, with widespread recovery by December 2020. The reduction in contraception-related activity (discussion and monitoring) was likely explained by a combination of reduced need (less social contact, use of non-prescription alternatives), longer repeat prescriptions, check-ups being postponed, and long acting reversible contraception (LARC), including coils and implants, not being fitted. Monthly contraceptive prescribing in England experienced only a small temporary reduction during the pandemic (^9^ and Figure S2). Six-week checks of infants were well maintained, likely prioritised as vital activities, possibly aided by increased use of telephone appointments and/or being carried out alongside 8-week immunisations, which were also prioritised.^30^ The slight increase in breast-related symptoms by December 2020 may indicate concerning delays in presentation. Current pregnancy records being slightly reduced may be explained by delayed presentation or increased use of self-referral directly to midwifery services. Other codes commonly recorded with pregnancy, e.g. date of last menstrual period, would be reduced for the same reasons. The low level of gestational diabetes indicates some codes for this condition were likely not captured here. Researchers have previously raised concerns about disruptions to sexual and reproductive health care services in the early stages of the pandemic.^31^ Although we observed a decline in female and reproductive health activities in primary care, most activities returned to near or above pre-pandemic levels by December.

#### Screening and Related Activity

Broadly, clinical activity related to screening declined substantially around April 2020, and there was widespread recovery by December to slightly above normal levels, with the exception of NHS health checks. The National Screening programme paused invitations for bowel screening in March 2020, and they were subsequently issued at rates above 100% of normal levels.^32^ NHS health checks were considered as “low priority” in the Royal College of General Practitioners (RCGP) workload prioritisation.^30^ The clinical advisory group did not propose any measures for ongoing monitoring.

Some studies outside the UK have investigated the impact caused by disruption to screening services, and found evidence for example that new breast cancer diagnoses were reduced^33^ and some groups may have been affected more than others.^34^

#### Medication Processes

Processes related to medication, in particular medication reviews, were relatively well maintained during the pandemic, likely due to automated alerts commonly prompting clinicians when these are due. Guidance on the new Structured Medicine Reviews was released in September 2020^35^ and uptake was relatively rapid. Other related codes, such as “Patient understands why taking all medication” are likely recorded during structured medication reviews, which explains why they also increased. Not all practices were recording Structured Medicine Reviews by January 2021, likely because pharmacists with the necessary training were not available in all practices. Use of this process is incentivised for 2021/22 for certain patient groups.^35^ The clinical advisory group proposed a key measure comprising any medication reviews.

### Policy implications and interpretation

The COVID-19 pandemic brought new challenges for the NHS to deliver safe and effective routine care. Despite the ongoing pressure of the pandemic, most primary care activities recovered to near-normal levels by December 2020, while some aspects of routine monitoring had not fully recovered. Our proposed *NHS Service Restoration Observatory* can support evaluation of national policies around service restoration and additionally provide opportunities for near real-time audit and feedback to rapidly identify and resolve concerns around health service activity. In particular we hope that data tools such as ours can be used to ensure continuity of high priority clinical services during subsequent waves of the pandemic.

### Future research

Across the clinical specialist areas we identified common themes for further research following this study with further detail on individual areas given (detailed list in Supplementary Information). OpenSAFELY is a national data resource and we encourage interested parties to consider exploring these patterns in the platform.

1. Monitoring activity in more granular groups such as those with established long term conditions, those receiving certain medicines, or those without prior history having new diagnoses, to establish the impact on each group and on new diagnoses vs ongoing monitoring.
2. The level of “backlog” could be analysed to inform NHS recovery plans and to establish whether those people who missed activities have later “caught up” and whether there are some groups waiting longer than others.
3. The pandemic offers an unprecedented natural experiment in new diagnoses and ongoing monitoring of patients’ conditions. Outcomes can be assessed to identify any clinical impacts on patients or tests that can be safely delayed without unintended impacts to free up health care capacity.
4. The impact on cancer referrals/stage at diagnosis in those with relevant symptoms e.g. breast symptoms or who missed screening.

5. Each topic should be assessed in the context of health inequalities to explore whether impacts affected some groups more than others, and should take into account other activity, e.g. prescriptions, referrals, and non-primary care activity.

#### Summary

We identified substantial and widespread changes in clinical activity in primary care since the onset of the COVID-19 pandemic, but there was generally good recovery by December 2020, with a few exceptions such as mental health which showed poor recovery, which warrant further investigation. The authors are now further developing the *OpenSAFELY NHS Service Restoration Observatory* for real-time monitoring of the key measures identified here as well as further studies, using primary care data to monitor and mitigate the indirect health impacts of Covid-19 on the NHS.

## Data Availability

All data produced are available online at https://github.com/opensafely/restoration-observatory-data-driven

https://github.com/opensafely/SRO-smr

## Administrative

## Acknowledgements

We are very grateful for all the support received from the TPP Technical Operations team throughout this work, and for generous assistance from the information governance and database teams at NHS England / NHSX. We also thank other contributors to the clinical advisory group including Raj Patel, Marion Wood, Rammya Mathew, and John Geddes. The views expressed are those of the authors and not necessarily those of the UK National Health Service, the NIHR, or the UK Department of Health.

## Information governance and ethical approval

NHS England is the data controller; TPP is the data processor; and the key researchers on OpenSAFELY are acting with the approval of NHS England. This implementation of OpenSAFELY is hosted within the TPP environment which is accredited to the ISO 27001 information security standard and is NHS IG Toolkit compliant;^36,37^ patient data has been pseudonymised for analysis and linkage using industry standard cryptographic hashing techniques; all pseudonymised datasets transmitted for linkage onto OpenSAFELY are encrypted; access to the platform is via a virtual private network (VPN) connection, restricted to a small group of researchers; the researchers hold contracts with NHS England and only access the platform to initiate database queries and statistical models; all database activity is logged; only aggregate statistical outputs leave the platform environment following best practice for anonymisation of results such as statistical disclosure control for low cell counts.^38^ The OpenSAFELY research platform adheres to the obligations of the UK General Data Protection Regulation (GDPR) and the Data Protection Act 2018. In March 2020, the Secretary of State for Health and Social Care used powers under the UK Health Service (Control of Patient Information) Regulations 2002 (COPI) to require organisations to process confidential patient information for the purposes of protecting public health, providing healthcare services to the public and monitoring and managing the COVID-19 outbreak and incidents of exposure; this sets aside the requirement for patient consent.^39^ Taken together, these provide the legal bases to link patient datasets on the OpenSAFELY platform. GP practices, from which the primary care data are obtained, are required to share relevant health information to support the public health response to the pandemic, and have been informed of the OpenSAFELY analytics platform.

This study was approved by the Health Research Authority (REC reference 20/LO/0651).

## Guarantor

HJC is guarantor.

