## Supplementary Information for "OpenSAFELY NHS Service Restoration Observatory 2: changes in primary care activity across six clinical areas during the COVID-19 pandemic"

**Methods** **2**
- Defining study measures 2
- Table S1. Definition of the study measures using CTV3 concepts and keywords. 3

Results **4**
- Table S2a. Cardiovascular disease codes 4
- Table S2b. Diabetes codes 5
- Table S2c. Mental Health codes 5
- Table S2d. Female and reproductive health codes 6
- Table S2e. Screening codes 7
- Table S2f. Medication Processes codes 8
- Figure S1. Antidepressant prescribing in primary care in England by month, Nov 2017 - Oct 2021 9
- Figure S2. Contraceptive prescribing in England by month, Nov 2017 - Oct 2021 10

Future research recommendations **10**
- CVD Further research 10
- Diabetes Further research 11
- Mental Health Further research 11
- Female and Reproductive Health Further research 11
- Screening Further research 11
- Medication processes Further research 12

### Methods

#### Defining study measures

For each topic, clinical codes were selected using two methods. First, we selected all CTV3 codes which were mapped to relevant CTV3 concepts ("cardiovascular" and "mental health disorder", Table S1). Second, we searched CTV3 code descriptions for pre-specified keywords to identify further clinical activities related to each topic of interest; this included searching descriptions associated with the parent CTV3 codes where codes were grouped. We checked the resulting lists of codes manually and excluded certain concepts, keywords or whole descriptions where required, to remove irrelevant items unintentionally captured by the keywords, to remove concepts not of interest (e.g. administrative processes or drug prescribing), or to remove codes with very low usage which did not produce interpretable decile charts (Table S1). We additionally looked at Structured Medication Reviews separately (https://github.com/opensafely/SRO-smr).

#### Table S1. Definition of the study measures using CTV3 concepts and keywords.

| **Measure** | | **CTV3 concept** | **Keywords** |
|---|---|---|---|
| **Cardiovascular Disease (CVD)** | Included | "cardiovascular" | "cardio" , "heart" , "cvd", "pulse", "blood pressure", "bp", "systolic", "diastolic" |
| | Excluded | (none) | "Other congenital heart anomalies", "(Cardiovascular procedures) or (Transfusions)", "(Cong heart dis, sept/bulb) or (bulbus cord) or (septal def)" |
| **Diabetes** | Included | (none) | "diabe", "DM", "insulin", "hypoglycaem", "desmond", "a1c" |
| | Excluded | "Drug" | (none) |
| **Mental health** | Included | "mental health disorder" | "mental", "learning", "dementia", "deleri", "psycho", "depress", "anxi", "cogn" |
| | Excluded | (none) | "environment" |
| **Female and reproductive health** | Included | (none) | "breast", "smear", "cervical", "contracept", "uterine", "iud", "coil", "cystosc", "preg", "female", "women", "matern", "vagi", "gynae", "obstet", "endomet", "fibroid", "hysterect", "hysterosc", "prolaps", "incontin" |
| | Excluded | "Administration", "(Neoplasms) or (cancers)" | "Complications of pregnancy,childbirth or the puerperium OS", "Gynaecological appliances", "Risk factors in pregnancy", "contraceptive implant", "non-obstetric" |
| **Screening and related procedures** | Included | (none) | "screen", "smear", or "NHS health check" |
| | Excluded | (none) | (none) |
| **Processes related to medication** | Included | (none) | "medication", "medicine", "drug", "presc", "repeat", or "rpt", or "structured medication reviews" |
| | Excluded | "Drug", "Causes of injury and poisoning", "Appliances+equipm ent" | "Supply of drugs payment admin", "Clinical trial administration (& drug)", "NHS 111 report received","OOH report","Telemedicine consultation" |

### Results

#### Table S2a. *Cardiovascular disease codes*

The most commonly recorded grouped and individual CTV3 codes in OpenSAFELY-TPP in 2020.

| | **CTV3 code** | **Description** | **2020 events (mill)** | **2020 Patient count (mill)** |
|---|---|---|---|---|
| High level grouping | 24 | Examination of cardiovascular system (& [vascu... | 27.17 | 6.55 |
| | 32 | Electrocardiography | 1.03 | 0.64 |
| | 18 | Cardiovascular symptoms (& [heart]) | 0.26 | 0.21 |
| | G5 | Other forms of heart disease | 0.18 | 0.13 |
| | G3 | Ischaemic heart disease (& [arteriosclerotic]) | 0.02 | 0.02 |
| | 79 | Heart procedure | 0.02 | 0.01 |
| | 54 | Contrast radiography excluding cardiovascular ... | 0.01 | 0.01 |
| Detailed codes | X773s | Pulse rate | 4.49 | 3.32 |
| | 243 | O/E - pulse rhythm (& [irregular]) | 1.66 | 1.40 |
| | XaQVY | QRISK2 cardiovascular disease 10 year risk score | 1.20 | 0.92 |
| | XM02J | Pulse regular | 1.20 | 1.01 |
| | 242 | O/E - pulse rate | 0.88 | 0.73 |
| | 24E | O/E periph pulses R-leg (& [dors ped][fem][pop... | 0.84 | 0.43 |
| | 24F | O/E periph pulses L leg (& [dors ped][fem][pop... | 0.84 | 0.43 |
| | XaKFx | Average home systolic blood pressure | 0.41 | 0.31 |
| | XaKFw | Average home diastolic blood pressure | 0.40 | 0.31 |

*O/E = on examination*

#### Table S2b. Diabetes codes

The most commonly recorded codes for diabetes activity by CTV3 code (January - December 2020).

| | CTV3 code | Description | 2020 events (mill) | 2020 Patient count (mill) |
|---|---|---|---|---|
| Detailed codes | XaPbt | Haemoglobin A1c level - IFCC standardised | 6.22 | 4.55 |
| | 66A | Diabetic monitoring | 1.47 | 0.66 |
| | X772q | Haemoglobin A1c level | 1.08 | 0.84 |
| | XaIeH | O/E - Right diabetic foot at low risk | 0.88 | 0.61 |
| | XaIeL | O/E - Left diabetic foot at low risk | 0.88 | 0.61 |
| | XaERp | HbA1c level (DCCT aligned) | 0.51 | 0.41 |
| | XaIlj | Diabetic retinopathy screening | 0.42 | 0.35 |
| | XaKSn | Diabetes care plan agreed | 0.34 | 0.26 |
| | 9OL | Diabetes administration: [monitoring] or [clinic] | 0.31 | 0.23 |
| | XaIuE | Diabetic foot examination | 0.24 | 0.23 |
| | X40J5 | Type II diabetes mellitus | 0.24 | 0.17 |

*O/E = on examination*

#### Table S2c. Mental Health codes

The most commonly recorded codes for mental health activity by CTV3 code (January - December 2020).

| | CTV3 code | Description | 2020 events (mill) | 2020 Patient count (mill) |
|---|---|---|---|---|
| High level grouping | E2 | Neurotic, personality and other nonpsychotic d... | 0.35 | 0.27 |
| | Eu | [X]Mental and behavioural disorders | 0.06 | 0.05 |
| | 28 | Nervous system and mental state general examin... | 0.06 | 0.05 |
| | 3A | Assessment: [mental disability] or [memory] or... | 0.05 | 0.03 |
| | E1 | Non-organic psychoses | 0.02 | 0.02 |
| | 9H | Mental health administration (& patient "secti... | 0.01 | 0.01 |
| | E | Mental health disorder | 0.01 | 0.01 |
| | 8G | Psychotherapy &/or sociotherapy | 0.01 | 0.01 |
| | d5 | Antipsychotic depot injections | 0.01 | <0.01 |
| | E0 | Organic psychotic condition | 0.01 | 0.01 |
| | Ez | Mental disorders NOS | <0.01 | <0.01 |
| | E3 | Mental retardation | <0.01 | <0.01 |

| | | | | |
|---|---|---|---|---|
| | d4 | Antipsychotic drug | <0.01 | <0.01 |
| | da | Other antidepressant drugs | <0.01 | <0.01 |
| Detailed codes | XaK6f | Depression interim review | 0.29 | 0.23 |
| | E20 | Neurotic disorder | 0.24 | 0.19 |
| | XE0re | Depressed mood | 0.22 | 0.18 |

*NOS = Not otherwise specified*

#### Table S2d. Female and reproductive health codes

*The most commonly recorded codes related to female and reproductive activity by CTV3 code (January - December 2020).*

| | **CTV3 code** | **Description** | **2020 events (mill)** | **2020 Patient count (mill)** |
|---|---|---|---|---|
| High level grouping | 61 | Contraception | 1.25 | 0.76 |
| | 15 | [Gynaecological] or [obstetric] history | 0.62 | 0.47 |
| | 64 | (Child health care)(inf feed meth)(breast/oth ... | 0.57 | 0.17 |
| | 62 | (Patient pregnant) or (pregnancy care [& anten... | 0.51 | 0.31 |
| | K5 | Other female genital tract disorders | 0.28 | 0.25 |
| | 27 | Obstetric examination | 0.18 | 0.04 |
| | 26 | Examination: [genitourinary] or [urinary] or [... | 0.12 | 0.11 |
| | K3 | Disorder of breast | 0.09 | 0.07 |
| | ga | Progesterone - only contraceptives | 0.08 | 0.04 |
| | 7E | Upper female genital tract operation | 0.07 | 0.06 |
| | L1 | Pregnancy complications | 0.06 | 0.04 |
| | 7F | Obstetric procedure (& puerperal) | 0.04 | 0.04 |
| | L0 | Pregnancy with abortive outcome | 0.04 | 0.03 |
| | 7D | Lower female genital tract operation | 0.02 | 0.01 |
| | 71 | Endocrine system &/or breast operations | 0.02 | 0.02 |
| | K4 | Female pelvic inflammatory disease | 0.01 | 0.01 |
| | gg | Vaginal lubricants | 0.01 | 0.01 |
| Detailed codes | Xa8Pl | Cervical smear | 1.11 | 1.06 |
| | XaKTi | Liquid based cervical cytology screening | 1.00 | 0.93 |
| | XaKd4 | Cervical cytology test | 0.84 | 0.44 |
| | XaPnn | Advice about long acting reversible contraception | 0.76 | 0.54 |
| | XaXfc | UK medical eligib criteria for contraceptive u... | 0.75 | 0.17 |
| | XE278 | Cervical smear - negative | 0.49 | 0.48 |

| | | | | |
|---|---|---|---|---|
| | 614 | Oral contraception | 0.43 | 0.28 |
| | 611 | General contraceptive advice | 0.43 | 0.32 |
| | XaagX | Cervical smear - human papillomavirus negative | 0.32 | 0.32 |
| | 621 | Patient currently pregnant | 0.32 | 0.22 |
| | X76Qu | Not pregnant | 0.27 | 0.21 |

#### Table S2e. Screening codes

The most commonly recorded codes related to screening activity by CTV3 code, January - December 2020, annotated with the screening type (bowel, cervical, diabetes, etc).

| CTV3 code | Description | 2020 events (mill) | 2020 Patient count (mill) | Screening type |
|---|---|---|---|---|
| XaPVj | Bowel cancer screening programme: faecal occul... | 1.29 | 1.28 | bowel |
| Y00e7 | Advice given - cervical smear | 1.12 | 0.56 | cervical |
| Xa8Pl | Cervical smear | 1.11 | 1.06 | cervical |
| XaKTi | Liquid based cervical cytology screening | 1.00 | 0.93 | cervical |
| Y0384 | Smear Under GMS | 0.99 | 0.97 | cervical |
| Y3562 | Smear consent given | 0.90 | 0.88 | cervical |
| Y0385 | Smear Slide Number | 0.83 | 0.82 | cervical |
| Y0390 | Smear Reason: Routine Recall | 0.66 | 0.66 | cervical |
| XE278 | Cervical smear - negative | 0.49 | 0.48 | cervical |
| XaIIj | Diabetic retinopathy screening | 0.42 | 0.35 | diabetes |
| 9O8 | Cervical smear screening administration | 0.41 | 0.32 | cervical |
| XaPf6 | No response to bowel cancer screening programm... | 0.40 | 0.37 | bowel |
| 68N | Immunisation status: [screen] or [consent status] | 0.33 | 0.27 | unspecified |
| XaagX | Cervical smear - human papillomavirus negative | 0.32 | 0.32 | cervical |
| XaRBT | NHS Health Check invitation first letter | 0.27 | 0.26 | health check |
| XaZJJ | Cervical smear screening SMS message text | 0.25 | 0.17 | cervical |
| Y0389 | Smear Reason: Routine Call | 0.24 | 0.24 | cervical |
| XaaPs | Human papillomavirus screening | 0.23 | 0.22 | cervical |
| Y0387 | Smear Clinical Data | 0.23 | 0.22 | cervical |
| XaMwb | Alcohol screen - AUDIT C completed | 0.21 | 0.20 | alcohol |
| XaRBQ | NHS Health Check completed | 0.21 | 0.18 | health check |

*GMS = General Medical Services*

#### Table S2f. Medication Processes codes

The most commonly recorded codes for processes related to medication by CTV3 code. (January - December 2020)

| CTV3 code | Description | 2020 events (mill) | 2020 Patient count (mill) |
|---|---|---|---|
| XaF8d | Medication review done | 7.25 | 5.12 |
| 8B3 | Drug therapy | 3.55 | 1.98 |
| XaZ08 | Has authorisation for medication under PSD | 2.48 | 2.06 |
| XaQA7 | Administration of medication under patient gro... | 1.81 | 1.49 |
| XaVvz | Administration of medication under patient spe... | 0.51 | 0.21 |
| XaJKW | Patient understands why taking all medication | 0.49 | 0.43 |
| XaIoW | Medication review done by pharmacist | 0.48 | 0.39 |
| Xa2yC | Able to manage medication | 0.40 | 0.37 |
| XaJKn | Drug compliance checked | 0.39 | 0.33 |
| XaIfK | Asthma medication review | 0.30 | 0.27 |
| XaJCO | Medication review with patient | 0.30 | 0.28 |
| XaJHr | Has shown no side effects from medication | 0.28 | 0.26 |
| XaMhk | Dispensing review of use of medicines | 0.23 | 0.20 |
| XaJCN | Medication review of medical notes | 0.23 | 0.18 |
| XaIVl | Medication review without patient | 0.23 | 0.17 |
| XaJJx | Indication for each drug checked | 0.23 | 0.21 |
| Y20a8 | Medication review done by clinical pharmacist | 0.22 | 0.18 |
| XaF8c | Medication review due | 0.22 | 0.16 |

*Codes related to patient group direction or patient specific direction (PSD) represent legal frameworks for administering medications such as vaccines.*

**Figure S1.** Antidepressant prescribing in primary care in England by month, Nov 2017 - Oct 2021

(source: https://openprescribing.net/bnf/0403/)

### 4.3: Antidepressant drugs

Part of chapter 4 Central Nervous System

High-level prescribing trends for Antidepressant drugs (BNF section 4.3) across all GP practices in NHS England for the last five years. You can explore prescribing trends for this section by CCG, or learn more about this site.

View all matching dm+d items.

#### Trends

Items Spending

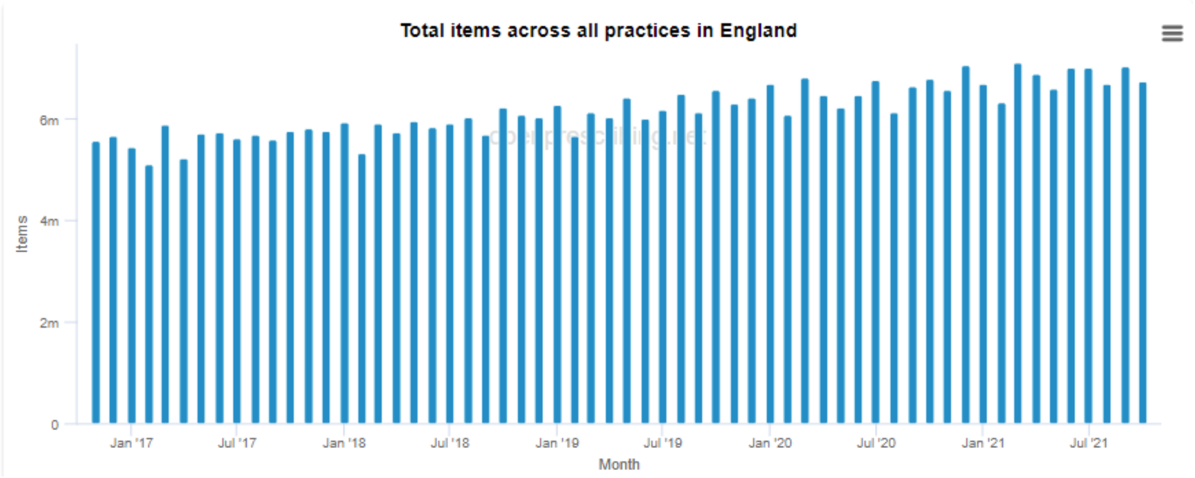

**Figure S2.** Contraceptive prescribing in England by month, Nov 2017 - Oct 2021

(source: https://openprescribing.net/bnf/0703/)

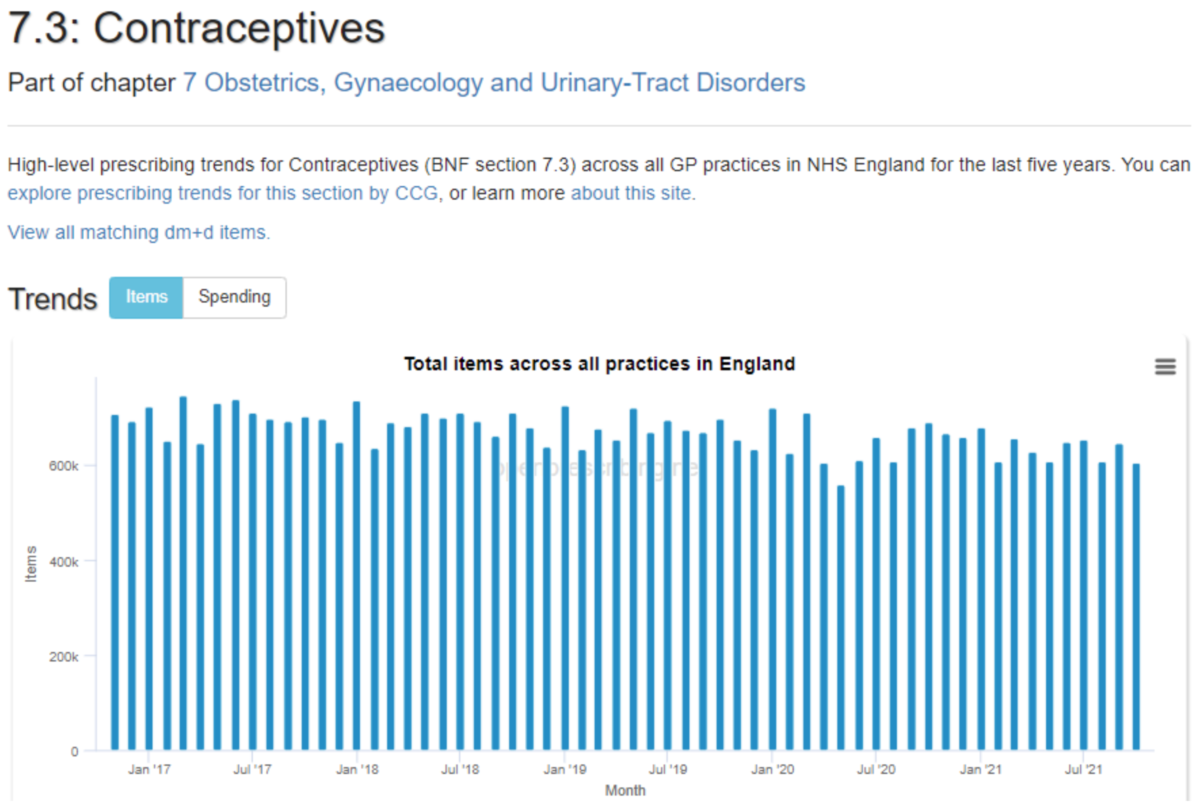


### Future research recommendations

#### CVD Further research

We identified several areas of interest for further research:

1. Codes could be analysed separately in people with and without prior diagnoses to establish the varying impacts on diagnosis and monitoring.
2. Blood pressure monitoring could be broken down into more granular groups such as those with established hypertension and those on contraceptives, to establish the impact on care of each group.
3. The level of "backlog" could be analysed to establish whether those people who missed out on routine monitoring have later had their appointment and whether there are some groups waiting long than others.

4. The level of clinical events such as hospitalisations could be measured to establish any impact of routine monitoring being reduced.

#### Diabetes Further research

We identified several areas of interest for further research:

1. The proportion of patients with diabetes receiving the appropriate monitoring and how this has been impacted by the pandemic.
2. The impact on the condition of those with diabetes could be investigated, for example whether HbA1C levels have generally worsened.
3. The rate of new diagnoses could be monitored.

#### Mental Health Further research

The potential for further research in this topic is limited due to the lack of access to data from mental health trusts and other services. However, a few areas were identified:

1. Additional conditions could be investigated such as personality disorders, eating disorders and ADHD.
2. Medications and histories could be used to give more context to the patterns of diagnosis coding.
3. The level of self harm incidents in hospital records (emergency attendances and admissions) could be investigated to establish any impact of routine monitoring or access being reduced. Similarly, alcohol misuse and associated violence-related injuries.

#### Female and Reproductive Health Further research

We identified several areas of interest for further research:

1. Impact on cancer referrals related to breast symptoms and bleeding.
2. Use of hormone-replacement therapy (HRT) and codes relating to the menopause.

#### Screening Further research

We identified several areas of interest for further research:

1. Cervical screening may be a potential candidate for ongoing monitoring.
2. The level of "backlog" could be analysed to establish whether those people who missed out on screening have later had their test and whether there are some groups waiting longer than others.
3. The rate of cancer diagnoses being made at more advanced stages could be investigated and any potential impact from missed screening (however, patients not choosing or not being able to consult for mild symptoms may also be a contributing factor).

#### Medication processes Further research

We identified several areas of interest for further research:

1. Medication reviews could be broken down into more granular groups such as those with dementia or asthma, to establish the impact on care of each group.
2. The level of "backlog" could be analysed to establish whether those people who missed out on routine monitoring have later had a review and whether there are some groups waiting longer than others.
3. The level of polypharmacy and adverse events associated with it (e.g. falls) could be measured to establish any impact of routine monitoring being reduced during the pandemic.
